# Experiences in the use of multiple doses of convalescent plasma in critically ill patients with COVID-19

**DOI:** 10.1101/2022.10.26.22278866

**Authors:** Ricardo Aguilar, Sandra López-Verges, Anarellys Quintana, Johanna Morris, Lineth Lopez, Ana Cooke, Dimas Quiel, Natalie Buitron, Yaseikiry Pérez, Lesbia Lobo, Maura Ballesteros, Yaneth Pitti, Yamilka Diaz, Lisseth Saenz, Danilo Franco, Daniel Castillo, Elimelec Valdespino, Isabel Blanco, Emilio Romero, Idalina Cubilla-Batista, Alcibiades Villarreal

**Affiliations:** Complejo Hospitalario Metropolitano Arnulfo Arias Madrid, Caja de Seguro Social, Panama, PANAMA; Hospital Santo Tomas, Panama, PANAMA; Hospital Punta Pacifica, Pacifica Salud, Panama, PANAMA; Hospital Rafael Estevez, Caja de Seguro Social, Aguadulce, PANAMA; Gorgas Memorial Institute of Health Studies, Panama, PANAMA; Sociedad Panameña de Hematología, Panama, PANAMA; Instituto de Investigaciones Científicas y Servicios de Alta Tecnología (INDICASAT-AIP), City of Knowledge, PANAMA; Universidad de Panama, PANAMA; Sistema Nacional de Investigación, SNI, SENACYT

**Keywords:** Convalescent plasma, severe acute respiratory syndrome, SARS-CoV-2, COVID-19, neutralizing antibodies

## Abstract

At the beginning of the SARS-CoV-2 pandemic, transfusion of COVID-19 convalescent plasma (CCP) was considered as one of the possibilities to help severe patients to overcome COVID-19 disease. The use of CCP has been controversial as its effectiveness depends on many variables from the plasma donor and the COVID-19 patient, for example, time of convalescence or symptoms onset. This was a feasibility study assessing the safety of multiple doses of CCP in mechanically ventilated intubated patients with respiratory failure due to COVID-19. Thirty (30) patients with severe respiratory failure, in ICU, with invasive mechanical ventilation received up to 5 doses of 300 to 600 ml of CP on alternate days (0,2,4,6 and 8) until extubation, futility, or death. Nineteen patients received five doses, seven received four, and four had 2 or 3 doses. On day 28 of follow-up, 57% of patients recovered and were at home and the long-term mortality observed was 27%. The ten severe adverse events reported in the study were unrelated to CCP transfusion. This study suggests that transfusion of multiple doses of convalescent plasma (CP) is safe. This strategy may represent an option to use in new studies, given the potential benefit of CCP transfusions in the early stage of infection in unvaccinated populations and in settings where monoclonal antibodies or antivirals are contraindicated or not available.

**Summary box:**

- Transfusion of multiple doses (up to 5 doses) of 300-600 ml of convalescent plasma from COVID-19 recovered patients is safe as it does not induce more severe effects than a single dose.
- Independent of the number of transfused doses, most patients had detectable levels of total and neutralizing antibodies in plasma.
- Future studies are needed to determine if multiple transfusion doses are more efficient in preventing severity than a single dose.

## Introduction

The Coronavirus disease 2019 (COVID-19) pandemic has led to an unexpected development of research in diagnosis and treatment areas (1). At the moment, with more than two years of the pandemic, the evidence-based treatment for COVID-19 patients of all types is rapidly changing to adjust accordingly. Not all antiviral therapies are well defined, and a few are still in authorization in many countries (2,3). There is evidence that not every patient with COVID-19 will benefit from passive immunotherapy (4,5), especially in patients with moderate and severe illness; clinical trials failed to find a reduction in mortality or improvement in clinical outcomes like mechanical ventilation use or length of hospital stay among others (6–9). Potentially, early transfused outpatients, seronegative, not vaccinated, or immunocompromised could benefit from the reduction in the risk for COVID-19 progression with convalescent plasma (CP) (10–12). Nevertheless, since COVID-19 convalescent plasma (CCP) is still described as an affordable therapeutic resource, clarification is warranted in preparation for future pandemic events (13). There is a need for improvement in the studies’ designs and methods to measure the outcomes (14,15), and there is still no clear definition of the optimal and safe CP dose and time to initiate transfusion nor the effectiveness of CP and monoclonal-antibody therapy in light of the SARS-CoV2 variants (4).

As part of the government’s strategies to ensure more options of treatments for COVID-19, at the beginning of the pandemic, in 2020, the Panamanian Society of Hematologists collaborated by launching a study to provide access to and evaluate the safety of use of multiple doses of CP in critically ill patients.

Any transfusion of plasma is associated with risks, including virus transmission (like HIV, HBV, HCV, and HTLV), allergic reactions, anaphylaxis, febrile reaction, transfusion-associated acute lung injury (TRALI), transfusion-associated circulatory overload (TACO), and hemolysis due to the administration of an incompatible ABO plasma unit (16–18).

This study aimed to assess whether the administration of multiple doses of CP was safe. Adverse events such as fever, rash, and infection related to the transfusion, TRALI, and TACO, were analyzed during the post-transfusion period of each CP unit administered. Other outcomes recorded were ventilatory-free days and mechanical ventilatory parameters, ICU (intensive care unit) time, hospital mortality, and long-term mortality by monitoring the 28 and 60-day mortality. Moreover, the antibody levels of donors and recipients were analyzed to study the possible association between the neutralizing immune response in the recipient with safety and survival.

## Methods

### Patients’ cohort and Bioethics’ considerations

This study was carried out at three hospitals in Panama City: Complejo Hospitalario Metropolitano Dr. Arnulfo Arias Madrid (CHMAAM) from the Social Security System, Hospital Santo Tomas, and a private hospital, Hospital Pacifica Salud, from June 1, 2020, through October 14, 2020. This study was conducted with the approval of the National Bioethics Committee (EC-CNBI-2020-04-56). The clinical research team informed every participant included in the study or a legally authorized representative about the study and the potential benefits and risks of using convalescent plasma as a therapy for critically ill patients.

### Inclusion and exclusion criteria of Convalescent Plasma recipients

We included patients aged ≥18 years old, hospitalized with COVID-19 severe respiratory symptoms in Intensive Care Unit (ICU) with invasive mechanical ventilation and with a positive laboratory diagnostic using SARS-CoV-2 RT-PCR test from nasopharyngeal swab. The exclusion criteria were contraindication to transfusion (severe volume overload, history of anaphylaxis to blood products), severe multi-organ failure, hemodynamic instability, other documented uncontrolled infections, severe Disseminated Intravascular Coagulation needing factor replacement, Fresh Frozen Plasma, cryoprecipitate, on dialysis, active intracranial bleeding or previous clinically significant myocardial ischemia.

### Data Collection

Clinical and laboratory (except antibodies measures) data were extracted from the patient’s medical record using a case report form designed for this study. We collected demographics, pre-existing medical conditions, COVID-19 symptoms, contagion routes, and timeline details. We followed the patients daily from one day before transfusion until death or extubation or day 27, whichever came first, and then, on days 28 and 60 from the first transfusion day, the team completed a follow-up call. The research team monitored for every patient during the transfusion the following criteria: daily vital signs, including SpO2, and daily ventilatory requirements such as: Fraction of inspired oxygen (FiO2), Positive End Expiratory Pressure (PEEP), and respiratory rate. New medical conditions, adverse events, evaluation for potential toxicities from CP transfusion were monitored and documented every 48 h (up to day 8). Data collection and management for this study was performed using ClinCapture, an open-source system, version 2.1.0. All categorical variables were described in frequencies or percentages and continuous variables were summarized with mean and standard deviation or median and the interquartile range (IQR). The data were analyzed using IBM SPSS Statistics (version 26, IBM Corp., 2019).

### Selection of Convalescent Plasma Donors

CP donors were individuals who had recovered from COVID-19 at least 14 days before the donation and had a negative SARS-CoV2 PCR test after recovery.

To recruit donors and achieve a sufficient CP stock for the study, the team balanced massive promotion with the information approved by the CNBI and personal interviews with potential donors. All donors provided written informed consent for donation (permit EC-CNBI-2020-04-44 for a first group of donors and EC-CNBI-2020-04-56 for the multiple donations) and samples were stored for further analysis.

Basic strategies to ensure the safety of donors and patients in the study were taken. Initially, only male or female donors with no history of pregnancy were accepted to minimize the possibility of TRALI. Subsequently, when the anti-HLA screening test was available, female donors with a history of pregnancy were allowed for CP donation (18). Suitable donors were evaluated for final screening at the Blood Bank from the CHMAAM. Every donation was screened for infections transmitted via transfusion (serology for HIV, HBV, HCV, HTLV-I/II, *T*.*cruzi, T*.*pallidum*, and Nucleic Acid Testing (NAT) methodology for HIV, HBV, and HCV).

An average of 600-700ml CP was extracted using Haemonetics, Trima, or Optia spectra apheresis, under surveillance of the medical staff and the investigators.

The CP units were inactivated by the psoralen methodology (19) and frozen as 200-250 ml aliquoted bags with their respective labels and stored at -70°C in a freezer set aside only for these units. The complete process of CP extraction included all quality controls required for other blood components. During this study, the Panamanian Ministry of Health issued an emergency use authorization of CP based on a similar approval from the FDA and convalescent plasma use in other pathologies.

### Convalescent Plasma Administration

Hospitalized subjects received open-label screened CP. Single or double plasma units (weight-based < or > 90Kg) were administered on days 0, 2, 4, 6, and 8, or until extubation or futility (if occurred before day 8) was determined by the ICU team. Treating clinicians could omit transfusion at their discretion (e.g., TRALI events are 100% donor-dependent and do not prohibit future transfusions) (20). All transfusions were ABO compatible. For each transfusion and every day between transfusions, the researchers assessed vital signs, physical condition status of ventilatory support, concomitant medication, and adverse event monitoring.

### Laboratory assays for SARS-CoV-2

For every subject, SARS-CoV-2 RNA presence was tested in nasopharyngeal swabs using RT-PCR before every plasma administration (21). Serum samples of donors and recipient patients were analyzed for the presence of anti-SARS COV-2 total IgG by ELISA (CE certified Vircell COVID-19 ELISA IgG, Vircell Spain S.L.U., Granada, Spain) according to the manufacturer’s recommendation. The ELISA assays use SARS-CoV-2 recombinant antigen from spike glycoprotein (S protein) and Nucleocapsid (N protein). To detect neutralizing antibodies against SARS-CoV-2 and determine their titer in plasma donor and recipient samples, samples were tested by performing a Plaque Reduction Neutralization Test (PRNT) using the SARS-CoV-2 virus original variant B.1. in Vero cells as previously described (22) in a BSL-3 laboratory. PRNT80 titer was calculated as the highest dilution of each sample capable of neutralizing the virus by inhibiting the formation of 80% of viral plaque units.

## Results

### Demographic and clinical description of the CP recipients during hospitalization

From June 5 until August 16 in 2020, a total of 30 patients were enrolled in this study. The majority of patients were male (67%), and the median age was 45.50 years (interquartile range (IQR)= 18). An 83% (25/30) of patients had at least one or more comorbidities. Obesity was the most prevalent chronic condition in 83%, followed by diabetes mellitus in 40% and hypertension in 40% of the patients. One patient had down syndrome, and one patient was in the second trimester of pregnancy. Five patients did not have comorbidities reported. Most patients (73%) had O-positive blood type. Thirty percent of the study subjects suspected exposure to SARS-CoV-2 was at work, 23% at home, 7% from their social network, and 40% did not know the route of exposure (Table 1).

**Table 1.**
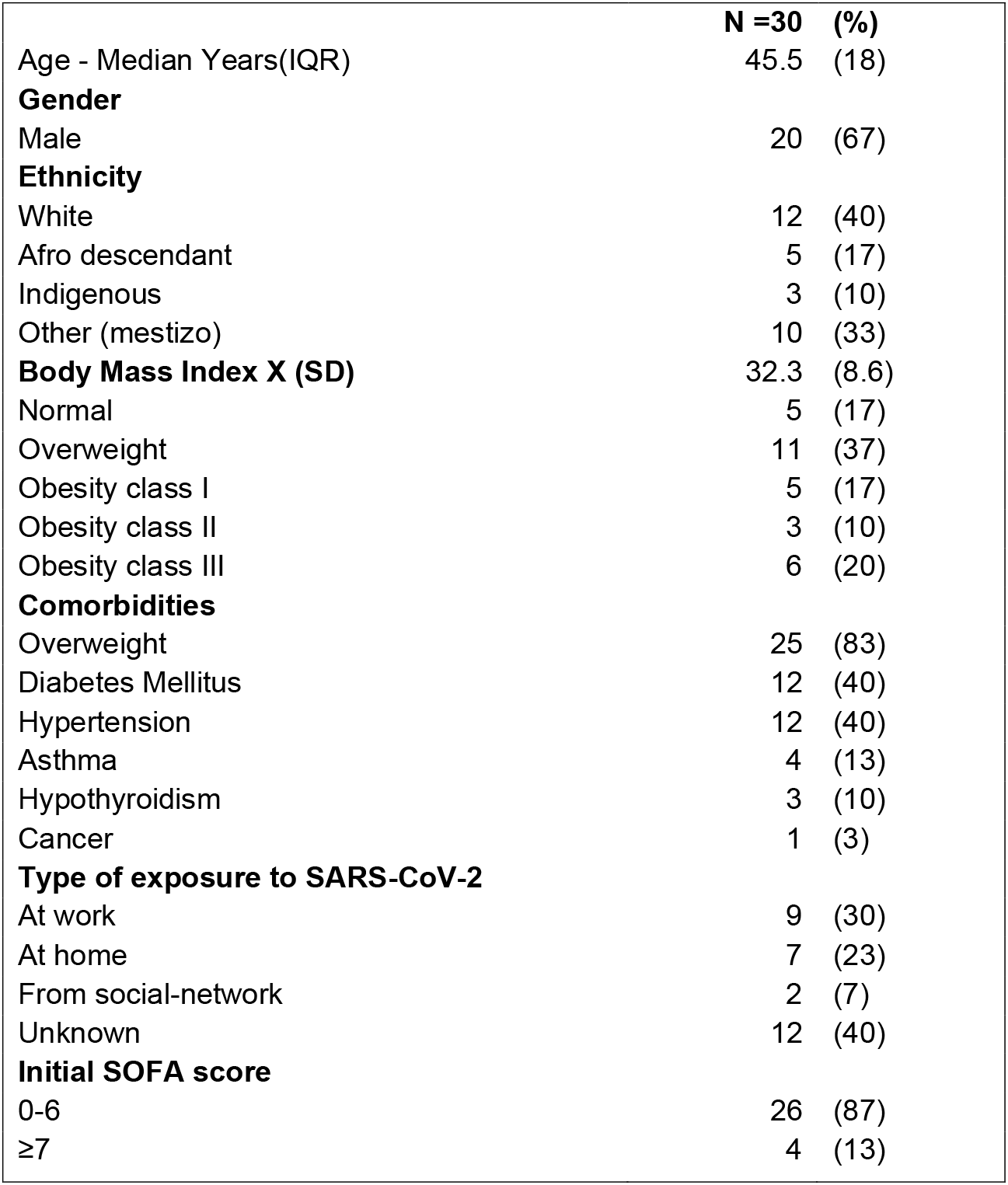
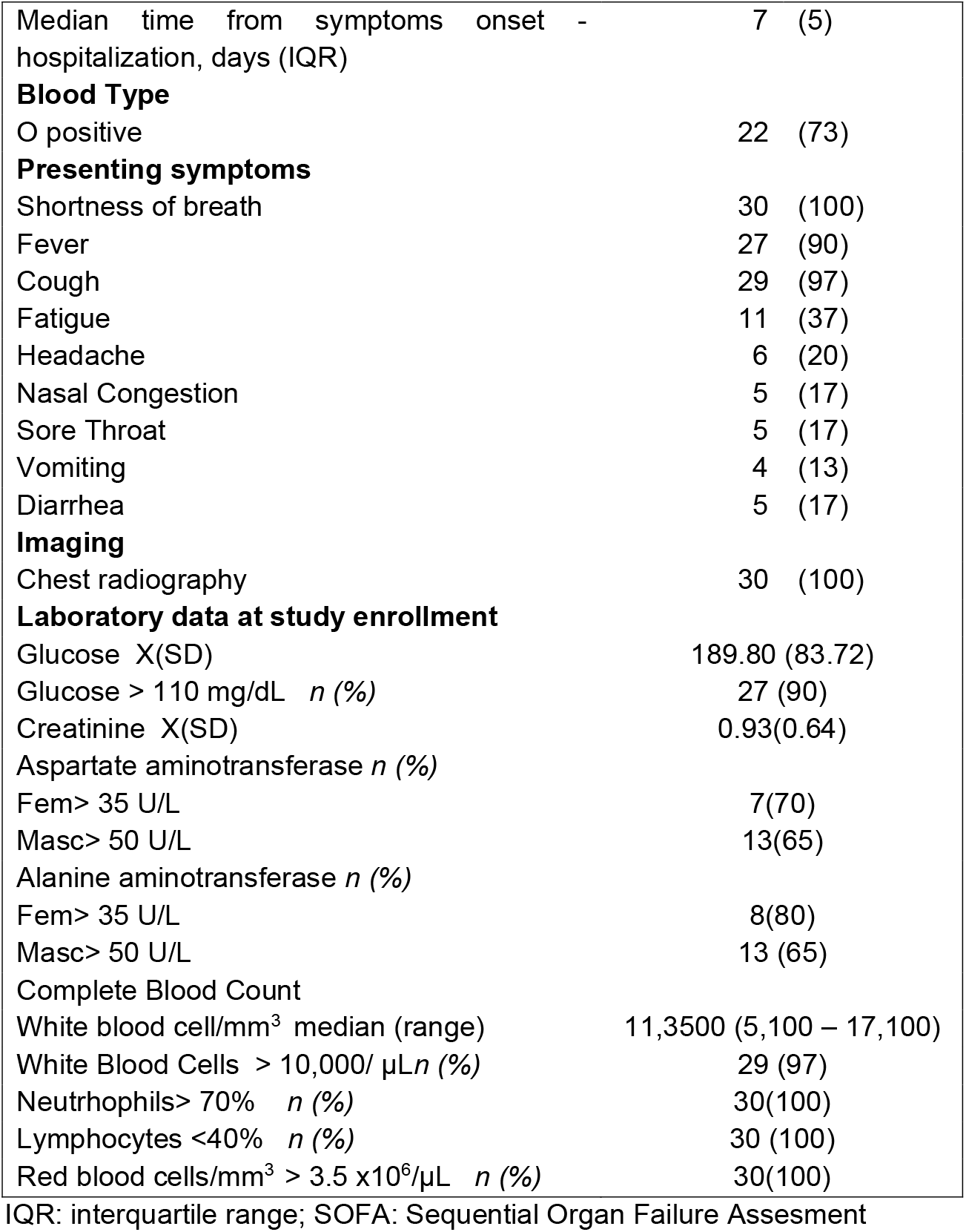
Characteristic of the convalescent plasma recipient sample cohort.

|  | **N =30** | **(%)** |
| --- | --- | --- |
| Age - Median Years(IQR) | 45.5 | (18) |
| **Gender** |  |  |
| Male | 20 | (67) |
| **Ethnicity** |  |  |
| White | 12 | (40) |
| Afro descendant | 5 | (17) |
| Indigenous | 3 | (10) |
| Other (mestizo) | 10 | (33) |
| **Body Mass Index X (SD)** | 32.3 | (8.6) |
| Normal | 5 | (17) |
| Overweight | 11 | (37) |
| Obesity class I | 5 | (17) |
| Obesity class II | 3 | (10) |
| Obesity class III | 6 | (20) |
| **Comorbidities** |  |  |
| Overweight | 25 | (83) |
| Diabetes Mellitus | 12 | (40) |
| Hypertension | 12 | (40) |
| Asthma | 4 | (13) |
| Hypothyroidism | 3 | (10) |
| Cancer | 1 | (3) |
| **Type of exposure to SARS-CoV-2** |  |  |
| At work | 9 | (30) |
| At home | 7 | (23) |
| From social-network | 2 | (7) |
| Unknown | 12 | (40) |
| **Initial SOFA score** |  |  |
| 0-6 | 26 | (87) |
| ≥7 | 4 | (13) |

|  |  |
| --- | --- |
| Median time from symptoms onset - hospitalization, days (IQR) | 7 (5) |
| **Blood Type** |  |
| O positive | 22 (73) |
| **Presenting symptoms** |  |
| Shortness of breath | 30 (100) |
| Fever | 27 (90) |
| Cough | 29 (97) |
| Fatigue | 11 (37) |
| Headache | 6 (20) |
| Nasal Congestion | 5 (17) |
| Sore Throat | 5 (17) |
| Vomiting | 4 (13) |
| Diarrhea | 5 (17) |
| **Imaging** |  |
| Chest radiography | 30 (100) |
| **Laboratory data at study enrollment** |  |
| Glucose X(SD) | 189.80 (83.72) |
| Glucose > 110 mg/dL *n* (%) | 27 (90) |
| Creatinine X(SD) | 0.93(0.64) |
| Aspartate aminotransferase *n* (%) |  |
| Fem> 35 U/L | 7(70) |
| Masc> 50 U/L | 13(65) |
| Alanine aminotransferase *n* (%) |  |
| Fem> 35 U/L | 8(80) |
| Masc> 50 U/L | 13 (65) |
| Complete Blood Count |  |
| White blood cell/mm<sup>3</sup> median (range) | 11,3500 (5,100 – 17,100) |
| White Blood Cells > 10,000/ μL *n* (%) | 29 (97) |
| Neutrophils> 70% *n* (%) | 30(100) |
| Lymphocytes <40% *n* (%) | 30 (100) |
| Red blood cells/mm<sup>3</sup> > 3.5 x10<sup>6</sup>/μL *n* (%) | 30(100) |

IQR: interquartile range; SOFA: Sequential Organ Failure Assessment

The median time from symptoms onset to hospitalization was seven days (IQR= 5 days) (Table 1). On the day of inclusion, every patient had a positive SARS-CoV2 RT-PCR test and most of them (87%) had an initial SOFA (Sequential Organ Failure Assessment) score equal or less than 6. The most common COVID-19 symptoms were shortness of breath, fever, and cough. Initial blood cell counts showed that most patients (97%) had a leukocytosis with marked lymphopenia before the first transfusion. Ninety percent of participants had an initial glucose level above 110 mg/dl. Among the most common medications administered were azithromycin (25/30), corticosteroids (28/30), heparin (29/30), and immunoglobulin (16/30) (Table 1). The median time from symptoms onset to the first plasma transfusion was 11 days (IQR=4 days), 16.5 days (IQR=29) for ICU hospitalization, 13 days (IQR=51) for the post-transfusion length of hospital stay until recovery or death, whereas the length of total hospital stay was a median of 21.5 days (IQR=54) (Table 2).

**Table 2.** Time intervals between symtopms onset, hospitalization, transfusion and final outcome including the drug treatment of convalescent plasma recipients during the hospitalization period.

| Patient No. | Time A | Time B | Drug treatments | Time C | Time D | Time E |
| --- | --- | --- | --- | --- | --- | --- |
| 1 | 15 | 1 | AZI,INM,COR,HEP | 15 | 14 | 13 |
| 2 | 16 | 6 | AZI,INM,COR,HEP | 24 | 24 | 10 |
| 3 | 12 | 3 | AZI,INM,COR,HEP | 56 | 33 | 43 |
| 4 | 8 | 1 | AZI,INM,COR,HEP | 20 | 16 | 8 |
| 5 | 15 | 2 | AZI, COR , HEP | 29 | 17 | 19 |
| 6 | 7 | 2 | AZI,INM,COR , HEP, VIT C, CEF | 11 | 6 | 2 |
| 7 | 10 | 2 | AZI,COR,HEP | 20 | 11 | 9 |
| 8 | 12 | 1 | AZI,INM,COR,HEP | 16 | 12 | 6 |
| 9 | 11 | 1 | AZI,INM,COR ,HEP | 15 | 8 | 11 |
| 10 | 11 | 2 | TOC,COR,HEP,IVER, VIT C, TIA | 37 | 17 | 28 |
| 11 | 11 | 1 | TOC,INM,HEP TEP,VITC, TIA | 17 | 17 | 16 |
| 12 | 7 | 1 | TOC,AZI,COR,HEP , TIA | 12 | 10 | 7 |
| 13 | 14 | 1 | AZI,INM,COR ,HEP | 18 | 15 | 14 |
| 14 | 8 | 2 | TOC,AZI,COR ,HEP TEP, | 28 | 11 | 18 |
| 15 | 10 | 2 | AZI,INM,COR ,HEP TEP, IVER | 59 | 35 | 49 |
| 16 | 12 | 2 | AZI,COR,HEP TEP,CEF | 23 | 20 | 12 |
| 17 | 14 | 2 | INM,COR ,HEP TEP | 31 | 26 | 24 |
| 18 | 14 | 4 | TOC,AZI,INM,COR ,HEP | 32 | 25 | 19 |
| 19 | 16 | 4 | AZI,COR,HEP , TIA, VIT C | 17 | 12 | 11 |
| 20 | 11 | 2 | TOC,COR,HEP TEP | 16 | 11 | 12 |
| 21 | 14 | 2 | COR ,HEP TEP | 55 | 28 | 40 |
| 22 | 9 | 2 | AZI,INM,COR,HEP TEP, VIT C, TIA, ZN, IVER | 15 | 15 | 13 |
| 23 | 10 | 3 | TOC,AZI,INM,HEP TEP, TAZ, IVER | 19 | 17 | 8 |
| 24 | 12 | 2 | AZI,INM,COR ,HEP TEP | 26 | 21 | 15 |
| 25 | 7 | 3 | HCQ,AZI,COR ,HEP TEP | 65 | 35 | 53 |
| 26 | 11 | 2 | COR INT,HEP,REM | 15 | 9 | 7 |
| 27 | 17 | 3 | AZI,COR ,HEP TEP | 24 | 14 | 12 |
| 28 | 15 | 3 | AZI,INM,COR ,HEP TEP | 27 | 19 | 18 |
| 29 | 12 | 2 | AZI,COR ,HEP | 34 | 27 | 25 |
| 30 | 10 | 2 | AZI,COR | 19 | 15 | 9 |
| **Median (IQR)** | **11.5 (4)** | **2 (1)** |  | **21.5 (15.3)** | **16.5 (13)** | **13 (11)** |

Time A: symptom onset to transfusion, days; Time B: Admission to ICU to transfusion, days; Time C: length of hospital stay, days; Time D: Length of ICU stay, days; Time E: Post-transf. length of hospital stay/death, days. AZI: Azithromycin; TOC: Tocilizumab; HCQ: Hydroxychloroquine; COR: corticosteroids; IVER: Ivermectin; HEP: prophylactic heparin; VIT C: Vitamin C; HEP TEP: therapeutic heparin; INM: immunoglobulin; TAZ: Tazobactam; REM: Resmidesivir; ZN:Zinc; TIA: Thiamine; CEF:Ceftriaxone

### Clinical Outcomes and safety profile of CCP transfusion

To monitor the clinical state of transfused patients, the oxygen support and clinical outcomes (7-points scale) was recorded before and after each transfusion (Figure 1A and Supplementary Figure 1). The percentage of extubated patients increased by day nine after the last CCP transfusion, 43% (13/30) of participants were out of mechanical ventilation and only 3.3% (1/30) had died.

**Figure 1.**
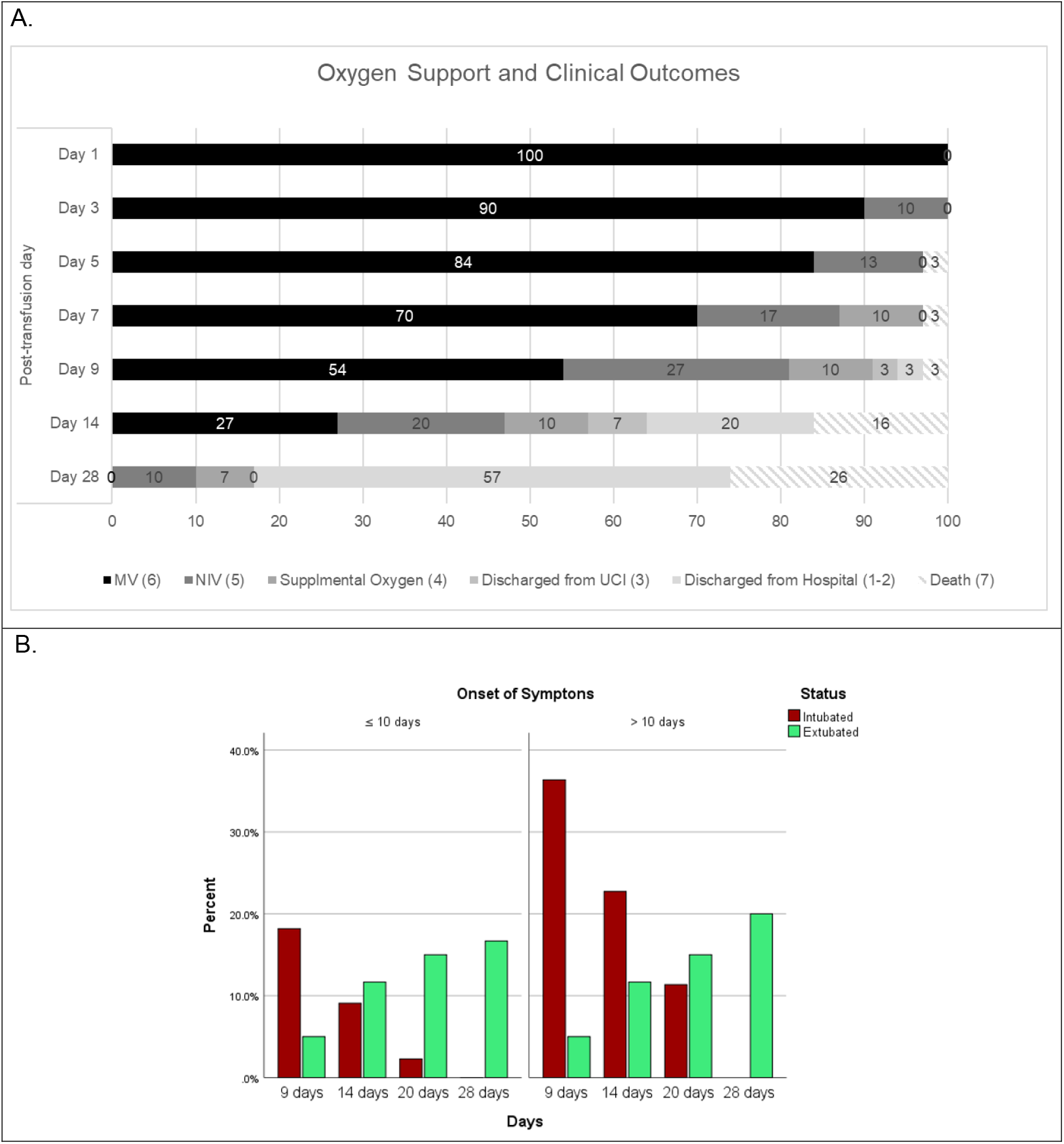
Clinical status and outcome of COVID-19 convalescent plasma transfusions. **A**. Graph showing the evolutions of patients in terms of oxygen support and 7-points scales for outcome (in parenthesis). MV: mechanical ventilation; NIV: non invasive ventilation. **B**. Oxygen support status (intubated or extubated) over time (post-transfusion days) depending on the time from symptoms onset to the the first CP transfusion (≤ 10 days or >10 days).

In this study, safety was the primary clinical endpoint. Even though the study proposed the transfusion of five doses of CP, the transfusion and the volume of the dose was contingent to extubation or medical condition. In twelve patients there was a reduction of the dose of CP transfused from 2 units (600 ml) to 1 unit (300 ml), due to their medical condition and in agreement with the ICU physician. Nineteen patients received five doses, seven patients received 4 doses and four patients were administered only 2 or 3 doses. Among the patients who received less than five doses, the major reason was extubation (ten patients), whereas one (1/30) died before receiving the complete proposed scheme. At day 28 of follow-up, 57% (17/30) of all patients recovered and were at home, 26% (8/30) died, the majority after 7 days were in ICU (Figure 1A). After 28 and 60 days of follow up the long-term mortality reported was 27%.

No adverse events attributed to plasma transfusion occurred in any case during the first 24 hours post-transfusion. Through the first eight days of follow-up during active transfusion, six moderate and one mild adverse event were registered, although none were related to CP transfusion. Among the moderate adverse events registered, one patient developed mild sinus bradycardia, and five patients developed hypotension, fever, gram-positive bacteremia, yeasts in tracheal secretions, and leg hematoma. None of the ten serious adverse events detected during the entire follow-up period was associated with the transfusion of CP. None of these adverse events precluded a reduction in the administration of doses of CP. Six serious adverse events happened in the first eight days of follow-up during active transfusion and four in 10-27 days of follow-up. Of these 10 serious adverse events, two patients that survived had sepsis secondary to bacteremia (A*chromobacter xylosoxidans, Staphylococcus epidermidis, Stenotrophomona maltophilia*). The other eight patients died, three due to multi-organ failure, one because of multi-organ failure and intractable pneumothorax, two due to septic shock, one because of sepsis plus intractable pneumothorax, and one due to respiratory failure.

### Presence of neutralizing antibodies and clinical outcome

Once safety multiple doses of CP transfusions were established, it was considered it was important to report if CP transfusion was associated or not with the development of neutralizing antibodies in recipient patients. At the moment of this study, it was considered that a COVID-19 convalescent patient with 14 days or more of symptoms resolution, and a SARS-CoV-2 negative PCR, could be a potential CP donor. For each of the five CP dose transfusions, the presence of total anti-SARS-CoV-2 IgG, and neutralizing activity was tested in donors and recipients. At the time of each transfusion, 90% to 100% of donors were positive for anti-SARS-CoV-2 IgG (Figure 2A and Supplementary Table 2). At day 0 (first transfusion) 90% of the CCP plasma units used had detectable levels of IgG, and 80% of the recipients also had detectable levels of IgG, showing no significant difference between recipients and the units transfused. Anti-SARS-CoV-2 IgG was detected in all recipients at day 4 post-transfusion. The neutralizing activity analysis of the CCP units used for the first transfusion showed that 43% had high neutralizing activity (PRNT_80_ titer ≥ 1:160), whereas 37% had a weak neutralizing activity (PRNT_80_ between 1:20 and 1:80) (Figure 2B and Supplementary Table 2). Mean days from symptom onset to donation were similar between donation with high neutralizing activity and low or no neutralizing activity (Supplementary Table 1). Although the difference of IgG detection in donors and recipient was not significant, almost 86% of recipients already had antibodies with neutralizing activity, 53% with strong neutralizing activity since first transfusion. The percentage of recipients having neutralizing antibodies increased overtime as expected, reaching 96% at day 4 after 2 transfusions (83% with strong neutralizing activity). However, the percentage of donors having neutralizing antibodies decreased overtime, reaching at day 8 of the study, only 21% with strong and around 64% with low neutralization titers. This prompted to compare the presence of IgG versus the neutralizing activity determined by PRNT_80_ in recipients before the first transfusion and at day 8. At day 0, only 3 recipients had no neutralizing antibodies even if most of them had detected IgG (Suplementary Figure 2). At day 8, all recipients had detected IgG with a high neutralization titer, only two had still a low neutralization activity. It appeared that there was no direct correlation between the IgG ELISA DO measurement and the neutralization titer calculated by PRNT_80_. Moreover, there was no association between IgG presence and neutralization titer before transfusion and post-transfusion with survival (Figure 2C and 2D).

**Figure 2.**
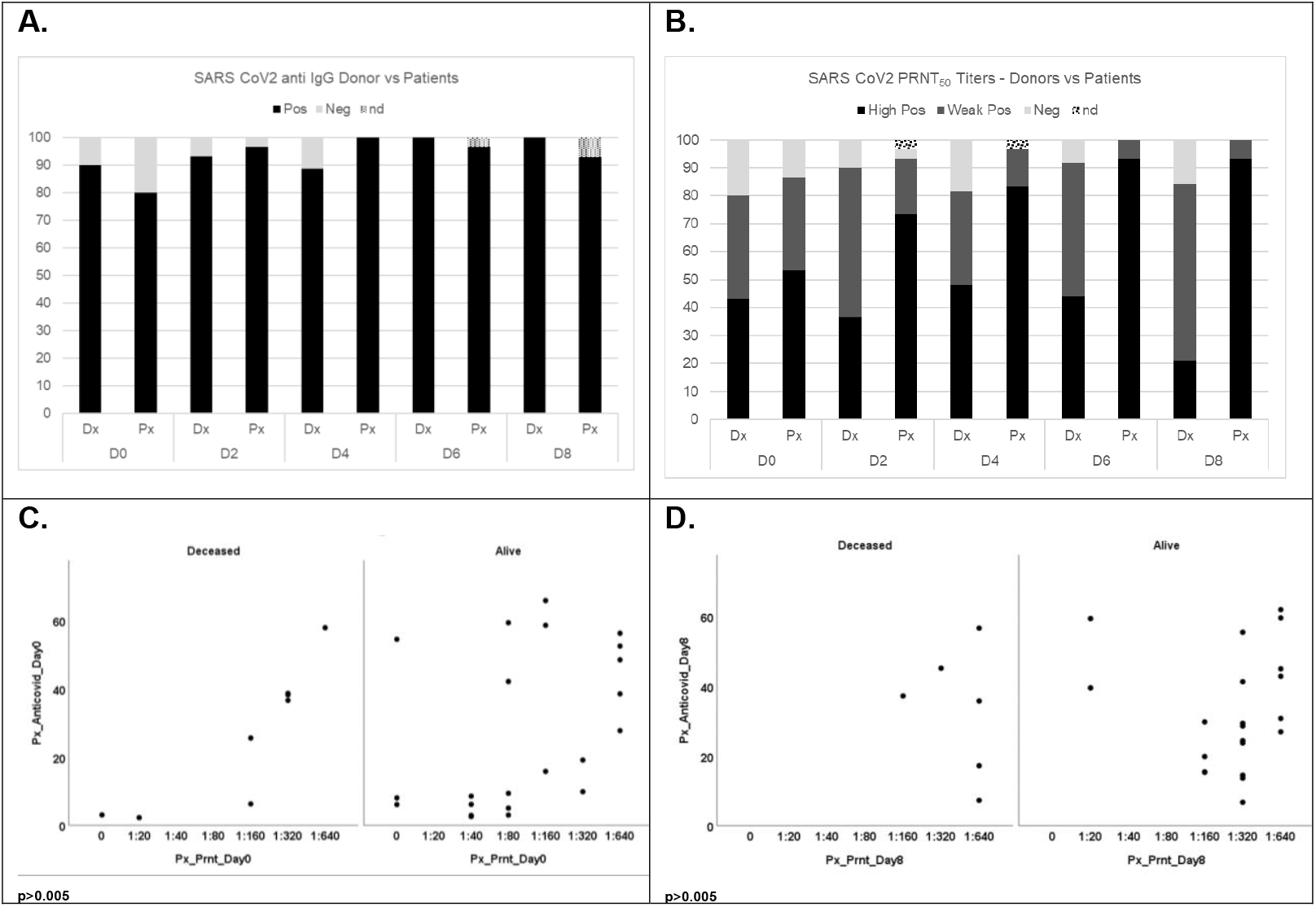
Detection of total IgG and neutralizing antibodies against SARS-CoV-2 for donors and recipients for each transfusion. **A**. Frequence of total anti-SARS-CoV-2 IgG in donors and recipient by day of transfusion (nd: no data)* **B**. Frequence of neutralization antibodies against SARS-CoV-2 in donors and recipient by day of transfusion. PRNT titers results are divided as follow: Negative=0; Low positive < 1:160; high positive ≥ 1:160.* **C and D**. Comparison of frequence of recipient patients positive for total anti-SARS-CoV-2 IgG versus neutralization level depending on the outcome (live or death). Comparison of total anti-SARS-CoV-2 IgG detection versus neutralization titer in recipient patients at day 0 before transfusion (**C**) and day 8 post-transfusion (**D**). *The total number of donors reported by days 4,6 and 8 were ≤30 since only 27, 25, and 20 patients were transfused respectively.

## Discussion

The primary endpoint of this trial was to assess the safety of multiple transfusions of convalescent plasma in critically ill patients, mainly allergic reactions, TACO or TRALI. Although we proposed the administration of more than 2 doses of convalescent plasma, there were no adverse reactions related nor volume overload, which is a sign of tolerability and safety. To our knowledge, this is one of the few studies where a volume ≥300ml was administered to a critically ill COVID-19 patient (11,23–25) with up to 5 doses in eight days, with no adverse events related to the CCP transfusion reported. These findings add useful information related to the therapeutic dose for overweight patients in pandemic, suggesting that transfusion of consecutive CCP units can be safe. In our study, the measures taken to ensure safety were likely the drivers to prevent this scenario. For example, the hematologist set up different strategies such as, the slow drip of the transfusions, the prescription of medication of patients with acetaminophen and diphenhydramine before the administration of CCP, and the careful search for specific donor characteristics as being a male, or female without previous pregnancies or screened negative for anti HLA antibodies. Another strategy to avoid overload in the patients was the reduction of the dose of plasma in agreement with the treating physicians when it was necessary. In patients with mechanical ventilation, the absence of adverse events related to the transfusions complement the evidence for safety of this treatment strategy.

Although this study was not designed to demonstrate efficacy, the difference in the proportion of extubated vs intubated patients depending on the time of transfusion after symptoms onset suggests a trend towards better outcomes when the transfusions were started earlier in the course of the disease. This is congruent with other reports that also observed that after day 9 of symptoms onset, the majority of patients already had potent neutralizing antibodies against SARS-CoV-2 previous to CP transfusion and with titers comparable to donors (10,12,24) and these titers increase or stay stable during the first month of follow up. Others showed the effectiveness of CP in early interventions (less than 7 days of symptoms onset) when there are still viral particles in the blood of the recipient that can be neutralized by the antibodies from the transfused plasma (26). The analysis of neutralization titers in recipients and donors allow us to confirm that transfusion should be done early when the recipients are still in the viremic phase and that donors should have more than the 14 days recovery as done in our study. Newer studies have shown than the stronger neutralizing antibody titers (27). After the analysis of our data, we agree with other authors that COVID-19 patients with high titers of IgG antibodies should not be transfused because it does not have an impact on outcomes and that the CP units used for transfusion should have high neutralizing antibody titers and be used early in the course of disease. We found that, 93% of participants received corticosteroids, thus it is possible that some of the improvement in the outcomes was related to the effect of this medication. Recent publications have stated the potential overlapping of benefits from both convalescent plasma and corticosteroid (28).

Even with new interventions as vaccines, COVID-19 remains a public health threat, especially when the SARS-Cov2 variants increase their circulation in every country. To the date seems that the CCP is more accessible than antivirals drugs and monoclonal antibodies principally in developing countries(28).

## Limitations

Not having the levels of IgG antibodies against SARS-Cov-2 in patients and donors at the time of transfusions was a major limitation of our study. In multiple cases, patients already had elevated levels of antibodies before being transfused and some donors had low levels of IgG. However, there is currently a different reality, as antibody tests are fully accessible. There are anti-RBD IgG antibody measurements, which have a very good correlation with neutralizing antibody levels (29).

Data extraction from clinical records it was not an easy task given the high number of cases and scarce information recorded per case at the beginning of the pandemic. Nevertheless, this study has an important value due to the data on safety of multiple doses of CP transfusion, as it served as a model, completed with the international data at that moment, for the national guidelines for convalescent plasma usage keeping the security measures, by extending it for compassionate use in the general population.

## Conclusion

This study has confirmed the safety profile of multiple COVID-19 convalescent plasma transfusions in critically ill patients, even if it did not demonstrate a benefit on disease outcome. This study could add evidence to support future clinical trials of the efficacy of multiple transfusions early in the course of the infection in patients who can’t be vaccinated, or can’t receive antivirals or monoclonal antibody treatments, like pregnant women or immunosuppressed patients; or in possible cases in which an emergent variant could escape the response of current vaccines preparations but not the neutralizing antibodies in the CP from survivors from this putative variant.

## Data Availability

All data produced in the present study are available upon reasonable request to the authors

---

**Supplementary Figure 1.**
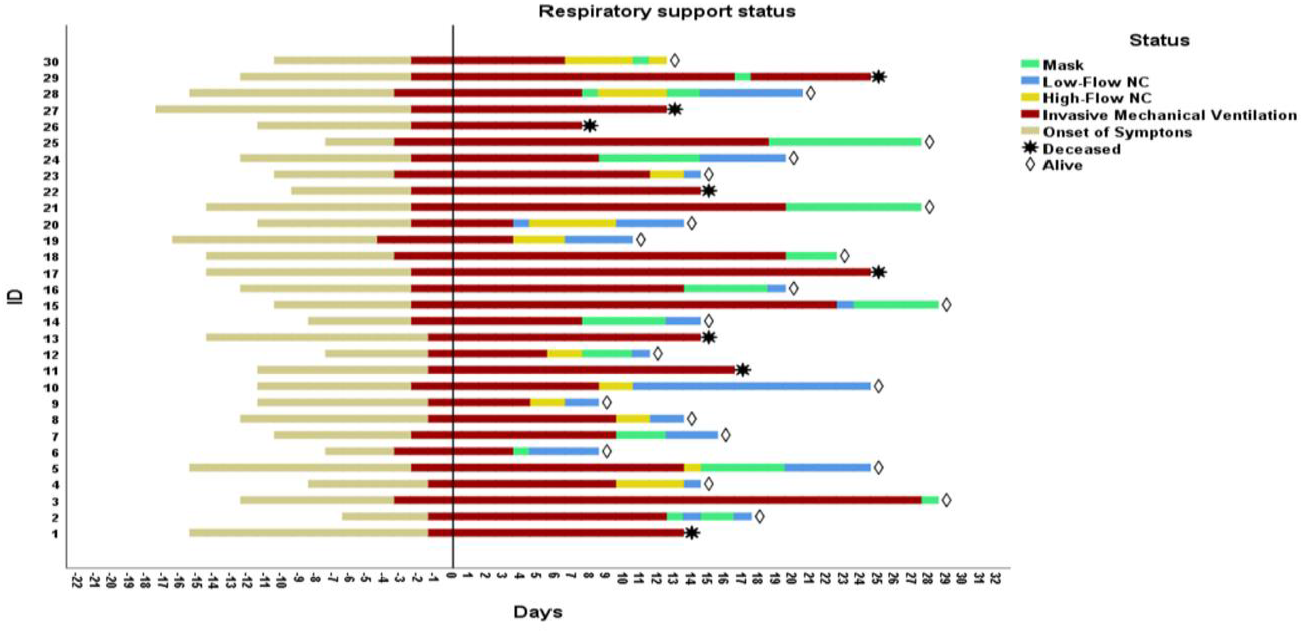
Oxygen support and clinical outcomes. Graph showing the onset of symptoms, the respiratory support status and the outcome (alive or death) for each CP recipient patient ID over time (in the timeframe, 0 corresponds to the day 0 of inclusion and first transfusion).

**Supplementary Figure 2.**
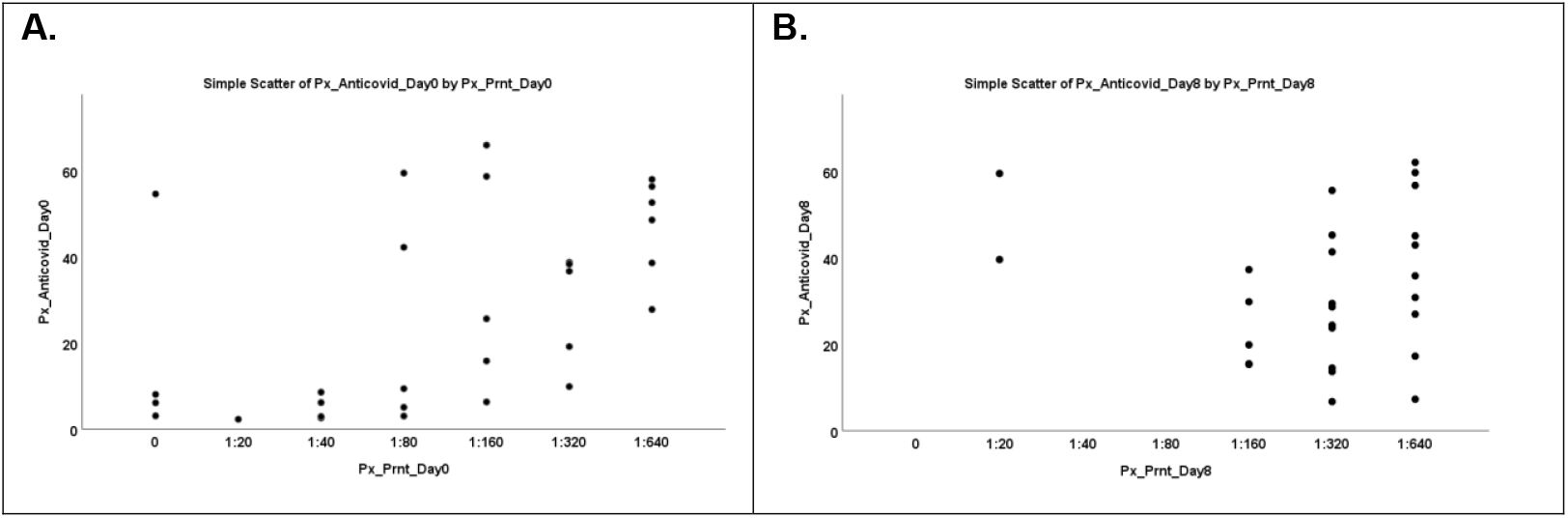
Comparison of frequence of recipient patients positive for total anti-SARS-CoV-2 IgG versus neutralization level by day of transfusion. Comparison of total anti-SARS-CoV-2 IgG detection versus neutralization titer in recipient patients at day 0 before transfusion (**A**) and day 8 post-transfusion (**D**).

**Supplementary Table 1.**
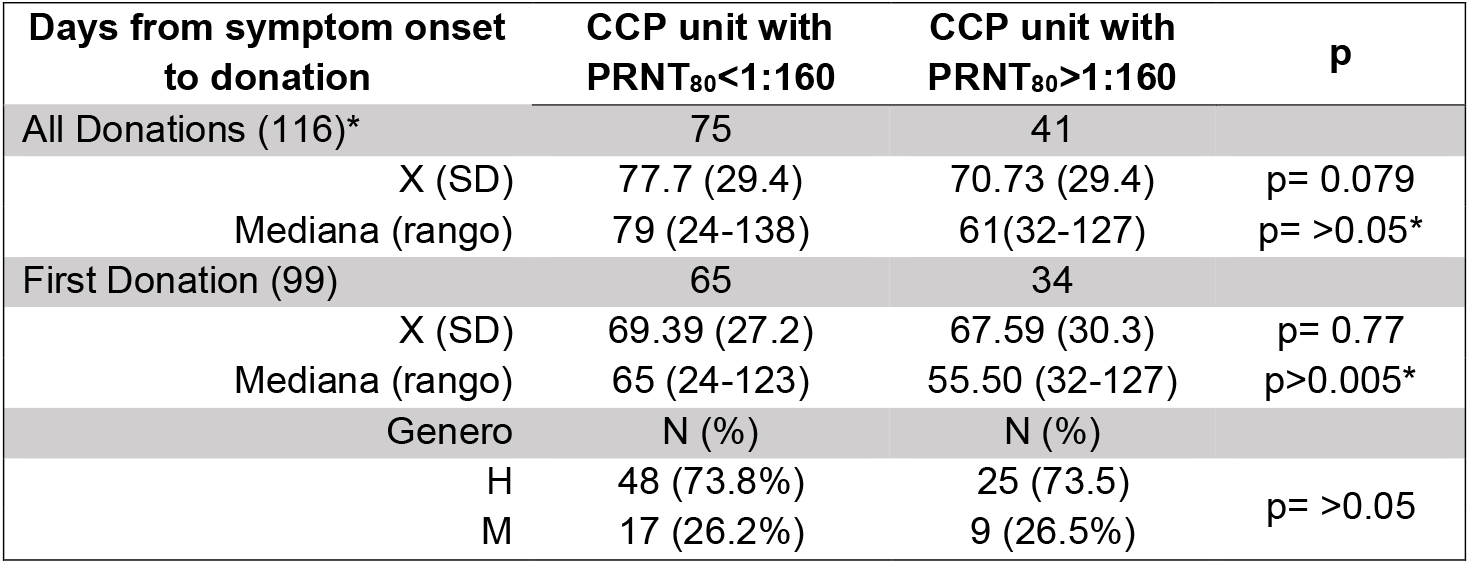
Summary of time between symptom onset and donation by COVID-19 Convalescent Plasma (CCP) unit with SARS-CoV2 neutralizing antibodies > 1:160. *Wilcoxon rank sum test.

| Days from symptom onset to donation | CCP unit with PRNT<sub>80</sub><1:160 | CCP unit with PRNT<sub>80</sub>>1:160 | p |
| --- | --- | --- | --- |
| All Donations (116)* | 75 | 41 |  |
| X (SD) | 77.7 (29.4) | 70.73 (29.4) | p= 0.079 |
| Mediana (rango) | 79 (24-138) | 61(32-127) | p= >0.05* |
| First Donation (99) | 65 | 34 |  |
| X (SD) | 69.39 (27.2) | 67.59 (30.3) | p= 0.77 |
| Mediana (rango) | 65 (24-123) | 55.50 (32-127) | p>0.005* |
| Genero | N (%) | N (%) |  |
| H | 48 (73.8%) | 25 (73.5) | p= >0.05 |
| M | 17 (26.2%) | 9 (26.5%) |  |

**Supplementary Table 2.**
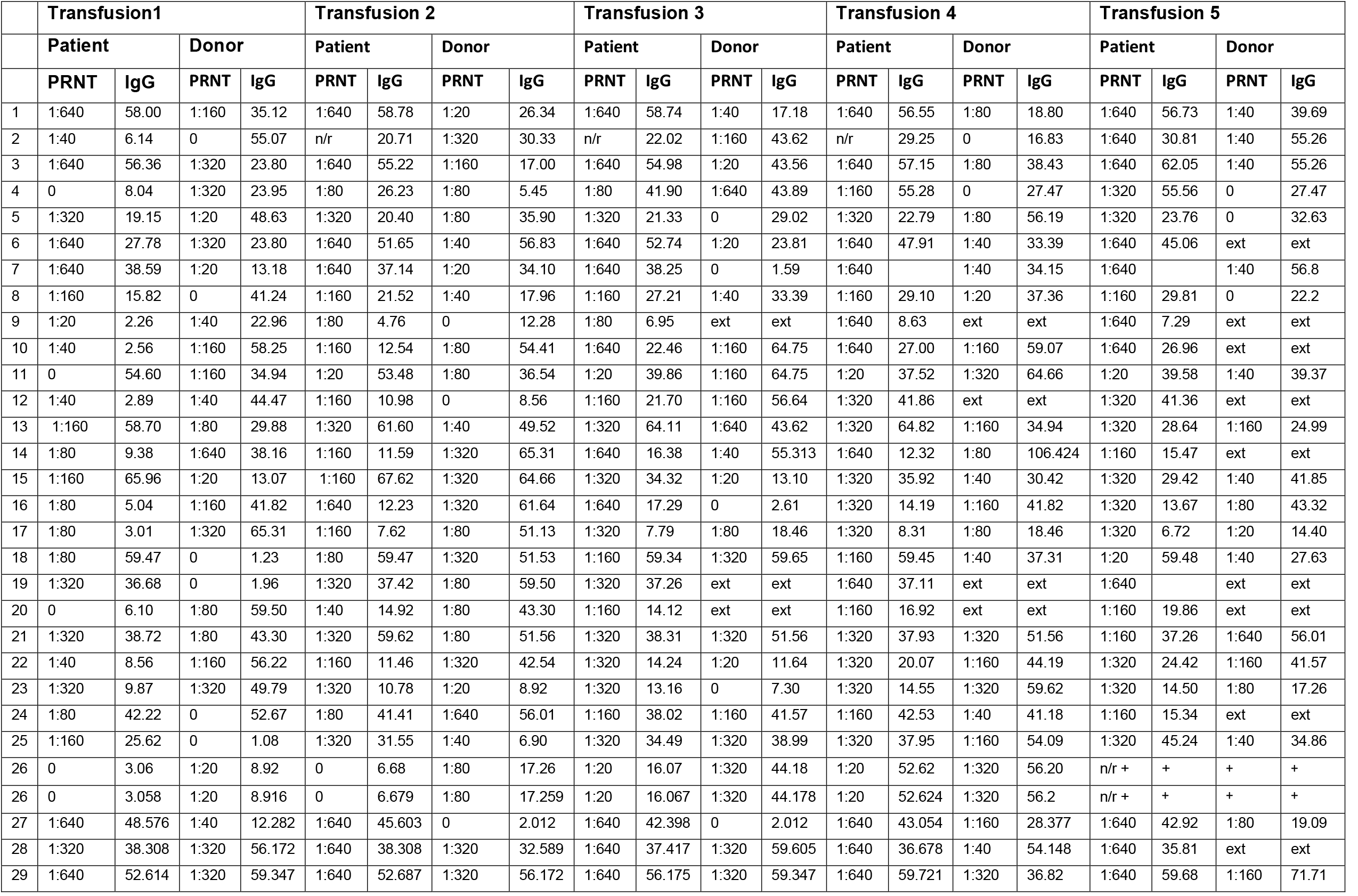
Presence of total anti-SARS-CoV-2 IgG and neutralizing antibodies in donors and recipients overtime. + Patient died; n/r: sample not received; ext: patient extubated

## Notes

### Competing Interest Statement

The authors have declared no competing interest.

### Clinical Trial

NCT05595538

### Funding Statement

This study was partially funded by Sistema Nacional de Investigacion

### Author Declarations

Ethics committee Comite Nacional de Bioetica/Panama gave ethical approval for this work. Protocol code EC-CNBI-2020-04-56 Approved

